# For-profit nursing homes and the risk of COVID-19 outbreaks and resident deaths in Ontario, Canada

**DOI:** 10.1101/2020.05.25.20112664

**Authors:** Nathan M. Stall, Aaron Jones, Kevin A. Brown, Paula A. Rochon, Andrew P. Costa

**Affiliations:** Division of General Internal Medicine and Geriatrics, Sinai Health System and the University Health Network, Toronto, Canada; Women’s College Research Institute, Women’s College Hospital, Toronto, Canada; Department of Medicine, University of Toronto, Toronto, Canada; Institute of Health Policy, Management and Evaluation, University of Toronto, Toronto, Canada; Department of Health Research Methods, Evidence, and Impact, McMaster University, Hamilton, Canada; Infection Prevention and Control, Public Health Ontario, Toronto, Canada; Dalla Lana School of Public Health, University of Toronto, Canada; Schlegel Chair in Clinical Epidemiology and Aging, McMaster University, Hamilton, Ontario, Canada; Centre for Integrated Care, St. Joseph’s Health System, Hamilton, Ontario, Canada

**Author notes:** **Corresponding author:** Nathan M. Stall MD, FRCPC, Division of General Internal Medicine and Geriatrics, Mount Sinai Hospital, Suite 475 - 600 University Avenue, Toronto ON M5G 2C4.

## Abstract

**Background:** Nursing homes have become the epicentre of the coronavirus disease 2019 (COVID-19) pandemic in Canada. Previous research demonstrates that for-profit nursing homes deliver inferior care across a variety of outcome and process measures, raising the question of whether for-profit homes have had worse COVID-19 outcomes than non-profit homes.

**Methods:** We conducted a retrospective cohort study of all nursing homes in Ontario, Canada from March 29-May 20, 2020 using a COVID-19 outbreak database maintained by the Ontario Ministry of Long-Term Care. We used hierarchical logistic and count-based methods to model the associations between nursing home profit status (for-profit, non-profit or municipal) and nursing home COVID-19 outbreaks, COVID-19 outbreak sizes, and COVID-19 resident deaths.

**Results:** The analysis included all 623 Ontario nursing homes, of which 360 (57.7%) were for-profit, 162 (26.0%) were non-profit, and 101 (16.2%) were municipal homes. There were 190 (30.5%) COVID-19 nursing home outbreaks involving 5218 residents (mean of 27.5 ± 41.3 residents per home), resulting in 1452 deaths (mean of 7.6 ± 12.7 residents per home) with an overall case fatality rate of 27.8%. The odds of a COVID-19 outbreak was associated with the incidence of COVID-19 in the health region surrounding a nursing home (adjusted odds ratio [aOR], 1.94; 95% confidence interval [CI] 1.23-3.09) and number of beds (aOR, 1.40; 95% CI 1.20-1.63), but not profit status. For-profit status was associated with both the size of a nursing home outbreak (adjusted risk ratio [aRR], 1.96; 95% CI 1.26-3.05) and the number of resident deaths (aRR, 1.78; 95% CI 1.03-3.07), compared to non-profit homes. These associations were mediated by a higher prevalence of older nursing home design standards in for-profit homes.

Interpretation: For-profit status is associated with the size of a COVID-19 nursing home outbreak and the number of resident deaths, but not the likelihood of outbreaks. Differences between for profit and non-profit homes are largely explained by older design standards, which should be a focus of infection control efforts and future policy.

## Introduction

Nursing homes have become the epicentre of the coronavirus disease 2019 (COVID-19) pandemic in Canada, with residents of these care homes accounting for more than 80% of the country’s deaths (1, 2). Nursing homes residents are at high risk of contracting severe acute respiratory syndrome coronavirus 2 virus (SARS-CoV-2) owing to their congregant living arrangement and exposure to staff with asymptomatic COVID-19 infection (3, 4). Nursing home residents are also at high risk of COVID-19 morbidity and mortality, with the majority being frail and multimorbid older adults (5). Despite these predisposing risks, there is widespread concern that nursing homes were both underprepared and underequipped to protect their residents, and many have questioned whether for-profit nursing homes have had worse COVID-19 outcomes (6, 7).

In Canada’s most populous province of Ontario, all nursing home residents receive personal and nursing care as well as subsidized accommodation under a publicly funded long-term care program. Regardless of this governmental funding, individual nursing homes can be owned and operated by for-profit, non-profit or public entities (8). Several observational studies suggest that for-profit nursing homes tend to deliver inferior care across a variety of outcome and process measures (9, 10). This includes lower levels and quality of staffing, more resident and family complaints, higher rates of emergency department visits, acute care hospitalizations and mortality (11-20). There is also evidence from the U.S. that for-profit nursing homes are more likely to receive deficiency citations for infection control and hand hygiene practices (21, 22). In light of this evidence and the catastrophic COVID-19 epidemic in nursing homes, we examined the association between for-profit status and the risk of COVID-19 outbreaks and death.

## Methods

### Study design

We conducted a retrospective cohort study of all 623 nursing homes in Canada’s most populous province of Ontario from March 29, 2020, the date of the first reported Ontario nursing home outbreak until May 20, 2020 (most recent data available) (23). The study was approved by the Research Ethics Board of the University of Toronto as well as the Hamilton Integrated Research Ethics Board. We followed the Strengthening the Reporting of Observational Studies in Epidemiology (STROBE) reporting guideline and the Reporting of Studies Conducted Using Observational Routinely-Collected Health Data (RECORD) statement guidelines (Appendix 1) (24, 25).

### Data sources

We obtained all data for this study from the Ontario Ministries of Health and Long-Term Care as part of the province’s emergency “modeling table”. This included nursing home level data from the Long-Term Care Inspections Branch on the cumulative number of resident COVID-19 cases and deaths. This data is collected daily by inspectors across the Province of Ontario who contact nursing homes and input data on outbreaks into a COVID-19 case tracking tool. The tracking tool also contains information on the profit status of the province’s nursing homes, which comprises for-profit homes (proprietary homes that are either owner operated or part of corporate chains), non-profit homes (charitable, religious and community agencies), and municipal public homes (municipally run and their employees are municipal government staff) (26).

Additional nursing home level data obtained from the Ontario Ministry of Long-Term Care included: number of licensed beds; the specific mix of bed occupancy types (one, two, or four residents per room); and the age of the nursing home’s design. The age of a nursing home’s design is determined by a structural classification of the home’s design standard (27). We classified nursing homes exceeding 1972 design standards as having ‘newer design standards’, and those homes meeting or falling below 1972 design standards as having ‘older design standards’. Homes with older design standards typically have smaller room sizes, less single occupancy rooms, and more shared washrooms (Appendix 2).

We also measured the cumulative incidence of COVID-19 in the communities surrounding each nursing home. This was calculated using deidentified line level data on all confirmed COVID-19 cases (as of May 19, 2020) from Ontario’s integrated Public Health Information System to determine the rate of COVID-19 per thousand individuals (using population data from the 2016 census) for each of Ontario’s 35 health regions served by a Public Health Unit (28). Residents of nursing homes were excluded from the numerator and denominator of this incidence calculation.

### Primary exposure and main outcomes

The primary exposure of interest was the nursing home profit status (for-profit, non-profit or municipal). The main outcomes of interest were: nursing home COVID-19 outbreaks (at least one resident case), COVID-19 outbreak sizes (total number of confirmed resident cases amongst homes with outbreaks), and total number of COVID-19 resident deaths (amongst homes with outbreaks).

### Statistical analysis

Summary statistics were computed to compare, by profit status, nursing home characteristics and the number of COVID-19 resident cases and deaths. We created separate multivariable statistical models for the three outcomes of interest. We modelled the risk of a nursing home COVID-19 outbreak with ≥1 resident case using logistic regression. COVID-19 outbreak size and total number of resident COVID-19 deaths were modelled using quasi-Poisson regression with an offset for the log of the number of beds within a nursing home. For all three models, random intercepts corresponding to each health region were included (29). For each of the three outcomes, we created three models all adjusted for the same factors. Model 1 included profit status only (unadjusted) and Model 2 included profit status + health region characteristics (population size of the nursing home location, COVID-19 cases per thousand in the health region surrounding the nursing home). Model 3 was an explanatory model and included profit status + health region characteristics + nursing home-level factors. We chose, *a priori*, to make our main inferences from model 2, and we used model 3 to examine explanatory nursing home factors related to profit status. We did not include bed occupancy type in the final models as this was collinear with the age of the nursing home design standard (Pearson correlation = 0.81). We examined the residual variability between health regions surrounding nursing homes using the median odds or risk ratio as well as percent change in variance (30). We also created dot charts to visualize the distribution of the size of COVID-19 outbreaks and the total number of deaths across nursing home profit statuses and age of design standards.

All covariates were selected *a priori* on the basis of a review of the literature. Analyses were performed using SAS statistical software, version 9.4 (SAS Institute Inc). Tests were 2-tailed, and the level of statistical significance was set at *α* = .05.

## Results

The analysis included all 623 Ontario nursing homes, of which 360 (57.7%) were for-profit homes, 162 (26.0%) were non-profit homes, and 101 (16.2%) were municipal homes. On average, and compared to both non-profit and municipal homes, for-profit homes were smaller (lowest mean number of licensed beds), had the lowest proportion of single occupancy rooms, and had older design standards (Table 1).

**Table 1:**
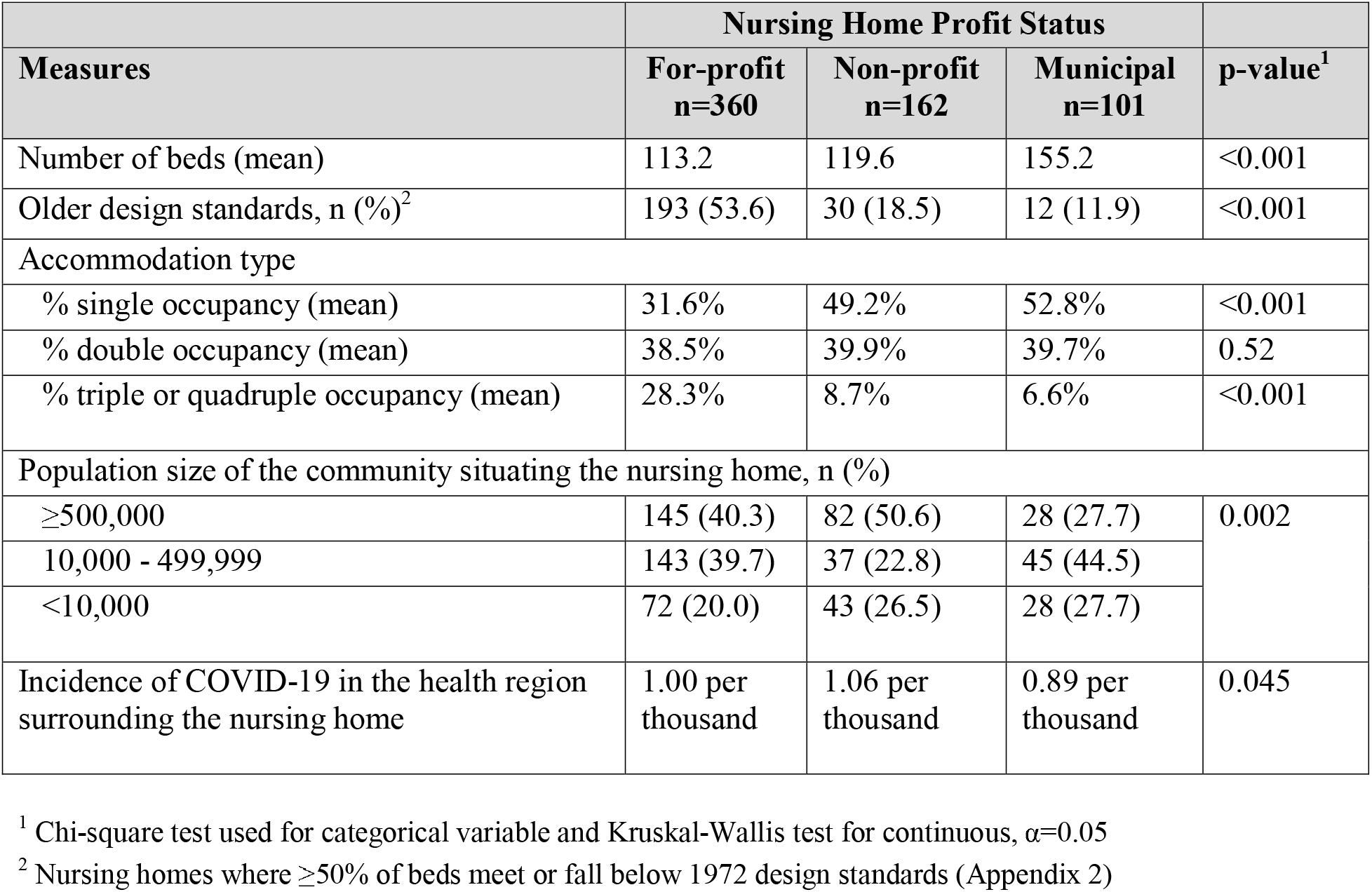
Characteristics of all 623 Ontario Nursing Homes by Profit Status.

| Measures | Nursing Home Profit Status |  |  | p-value<sup>1</sup> |
| --- | --- | --- | --- | --- |
|  | For-profit<br>n=360 | Non-profit<br>n=162 | Municipal<br>n=101 |  |
| Number of beds (mean) | 113.2 | 119.6 | 155.2 | <0.001 |
| Older design standards, n (%)<sup>2</sup> | 193 (53.6) | 30 (18.5) | 12 (11.9) | <0.001 |
| Accommodation type |  |  |  |  |
| % single occupancy (mean) | 31.6% | 49.2% | 52.8% | <0.001 |
| % double occupancy (mean) | 38.5% | 39.9% | 39.7% | 0.52 |
| % triple or quadruple occupancy (mean) | 28.3% | 8.7% | 6.6% | <0.001 |
| Population size of the community situating the nursing home, n (%) |  |  |  |  |
| ≥500,000 | 145 (40.3) | 82 (50.6) | 28 (27.7) | 0.002 |
| 10,000 - 499,999 | 143 (39.7) | 37 (22.8) | 45 (44.5) |  |
| <10,000 | 72 (20.0) | 43 (26.5) | 28 (27.7) |  |
| Incidence of COVID-19 in the health region surrounding the nursing home | 1.00 per thousand | 1.06 per thousand | 0.89 per thousand | 0.045 |

<sup>1</sup> Chi-square test used for categorical variable and Kruskal-Wallis test for continuous, $\alpha=0.05$
<sup>2</sup> Nursing homes where ≥50% of beds meet or fall below 1972 design standards (Appendix 2)

Overall, the crude incidence of COVID-19 nursing home outbreaks was 85.1 per thousand among for-profit homes, 61.4 per thousand among non-profit homes, and 23.4 per thousand among municipal homes. The crude rate of COVID-19 nursing home resident deaths was 23.4 per thousand among for-profit homes, 18.2 per thousand among non-profit homes, and 5.8 per thousand among municipal homes. The case-fatality rate among nursing home residents was 27.5% among for-profit homes, 29.7% among non-profit homes, and 25.0% among municipal homes (Table 2).

**Table 2:** Ontario Nursing Home COVID-19 Outbreaks and Deaths by Profit Status (March 29-May 20, 2020)

| Measures | Nursing Home Profit Status |  |  | p-value<sup>1</sup> |
| --- | --- | --- | --- | --- |
|  | For-profit<br>n=360 | Non-profit<br>n=162 | Municipal<br>n=101 |  |
| COVID-19 Outbreaks |  |  |  |  |
| Total number of COVID-19 cases | 3599 | 1239 | 380 | - |
| Incidence of COVID-19 cases | 85.1 per<br>thousand | 61.4 per<br>thousand | 23.4 per<br>thousand | <0.001 |
| Homes with a resident outbreak, n (%) | 110 (30.6) | 55 (34.0) | 25 (24.8) | 0.29 |
| Proportion of residents infected per outbreak<br>home, mean (SD) | 23.8%<br>(.303) | 17.2%<br>(.215) | 7.1% (.129) | 0.01 |
| Mean number of cases per outbreak home (SD) | 32.7 (48.0) | 22.5 (26.7) | 15.2 (30.1) | 0.20 |
| COVID-19 Resident Deaths |  |  |  |  |
| Total number of COVID-19 deaths | 989 | 368 | 95 | - |
| COVID-19 death rate | 23.4 per<br>thousand | 18.2 per<br>thousand | 5.8 per<br>thousand | <0.001 |
| Homes with any resident death, n (%) | 51 (14) | 33 (20) | 11 (11) | 0.086 |
| Proportion of resident deaths per outbreak home,<br>mean (SD) | 6.5%<br>(.101) | 5.5%<br>(0.081) | 1.7%<br>(0.037) | 0.19 |
| Mean number of deaths per outbreak home (SD) | 9.0 (14.6) | 6.7 (9.4) | 3.8 (8.8) | 0.33 |
| Case fatality rate % | 27.5% | 29.7% | 25.0% | 0.14 |

<sup>1</sup> Exact binomial test used for incidence, chi-square test for categorical variables, and Kruskal-Wallis test for continuous, $\alpha=0.05$

### Odds of a COVID-19 nursing home outbreak

There were 190 (30.5%) COVID-19 outbreaks among Ontario’s nursing homes, with 110 (30.6%) occurring in for-profit homes, 55 (34.0%) occurring in non-profit homes and 25 (24.8%) occurring in municipal homes (Table 2). In the unadjusted (model 1) and health region characteristics-adjusted (model 2) logistic regression models, nursing home profit status was not significantly associated with the odds of a COVID-19 nursing home outbreak (Table 3). In the fully adjusted explanatory model (model 3), both the incidence of COVID-19 in the health region surrounding the nursing home (adjusted odds ratio [aOR], 1.94; 95% confidence interval [CI] 1.23-3.09; per increase in 1/1000 COVID-19 incidence) and the total number of beds (aOR, 1.40; 95% CI 1.20-1.63; per 50 beds) were significantly associated with the odds of a COVID-19 nursing home outbreak.

**Table 3:** Odds of a Nursing Home COVID-19 Outbreak by Profit Status.

|  | **Model 1**<br>(Profit status only) | **Model 2**<br>(+ Adjustment for<br>health region<br>characteristics) | **Model 3 (Explanatory)**<br>(+ Adjustment for<br>nursing home<br>characteristics) |
| --- | --- | --- | --- |
| **Variable** | **aOR (95% CI)** | **aOR (95% CI)** | **aOR (95% CI)** |
| **Profit status** |  |  |  |
| Non-profit (ref) | - | - | - |
| For-profit | 1.01 (0.64-1.57) | 0.96 (0.61-1.49) | 0.88 (0.55-1.41) |
| Municipal | 0.83 (0.45-1.54) | 0.85 (0.46-1.58) | 0.64 (0.23-1.23) |
| **Health region characteristics** |  |  |  |
| COVID-19 incidence<br>(1 case per thousand) |  | 2.02 (1.20-3.38) | 1.94 (1.23-3.09) |
| Population |  |  |  |
| >500,000 (ref) |  | - | - |
| 10,000 - 499,999 |  | 0.57 (0.32-1.00) | 0.56 (0.33-0.94) |
| <10,000 |  | 0.27 (0.13-0.58) | 0.40 (0.19-0.85) |
| **Nursing home characteristics** |  |  |  |
| Number of beds (unit of<br>50) |  |  | 1.40 (1.20-1.63) |
| Older design standards |  |  | 1.48 (0.97-2.25) |

aOR = adjusted odds ratio

### Size of COVID-19 nursing home outbreaks (total number of resident cases)

Among nursing homes with a COVID-19 outbreak, an average of 23.8% of all residents in for-profit homes had COVID-19, whereas on average 17.2% and 7.1% of all residents in non-profit and municipal homes had COVID-19, respectively (Table 2). Thirteen of the fifteen homes with the highest infection rates were for-profit-homes with older design standards (Figure 1). In both unadjusted (risk ratio [RR], 1.83; 95% CI 1.18-2.84) and health region characteristics-adjusted quasi-Poisson regression models (adjusted RR [aRR], 1.96; 95% CI 1.26-3.05), for-profit status was significantly associated with the size of the COVID-19 nursing home outbreak compared to non-profit status (Table 4). In the fully adjusted explanatory model, the relationship with for-profit status was attenuated (aRR, 1.35; 95% CI 0.86-2.12), whereas both the number of COVID-19 cases per thousand in the health region surrounding the nursing home (aRR, 1.70; 95% CI 1.04-2.76), older nursing home design standards (aRR, 1.73; 95% CI 1.17-2.56), and total number of beds (aRR, 0.82; 95% CI 0.72-0.93; per 50 beds) were significantly associated with the size a COVID-19 nursing home outbreak.

**Figure 1:**
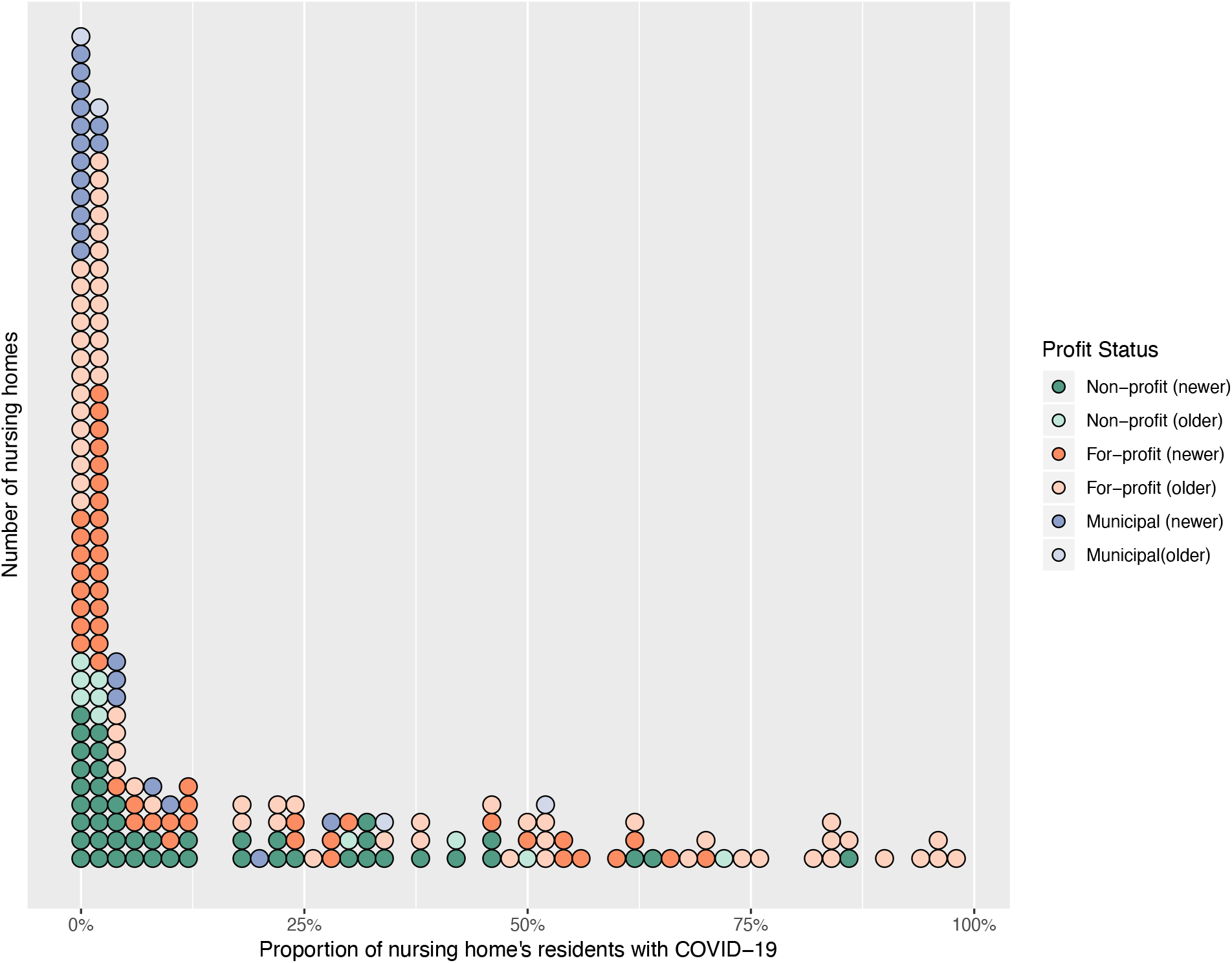
Distribution of Nursing Home COVID-19 Outbreak Sizes by Profit Status.

**Table 4:**
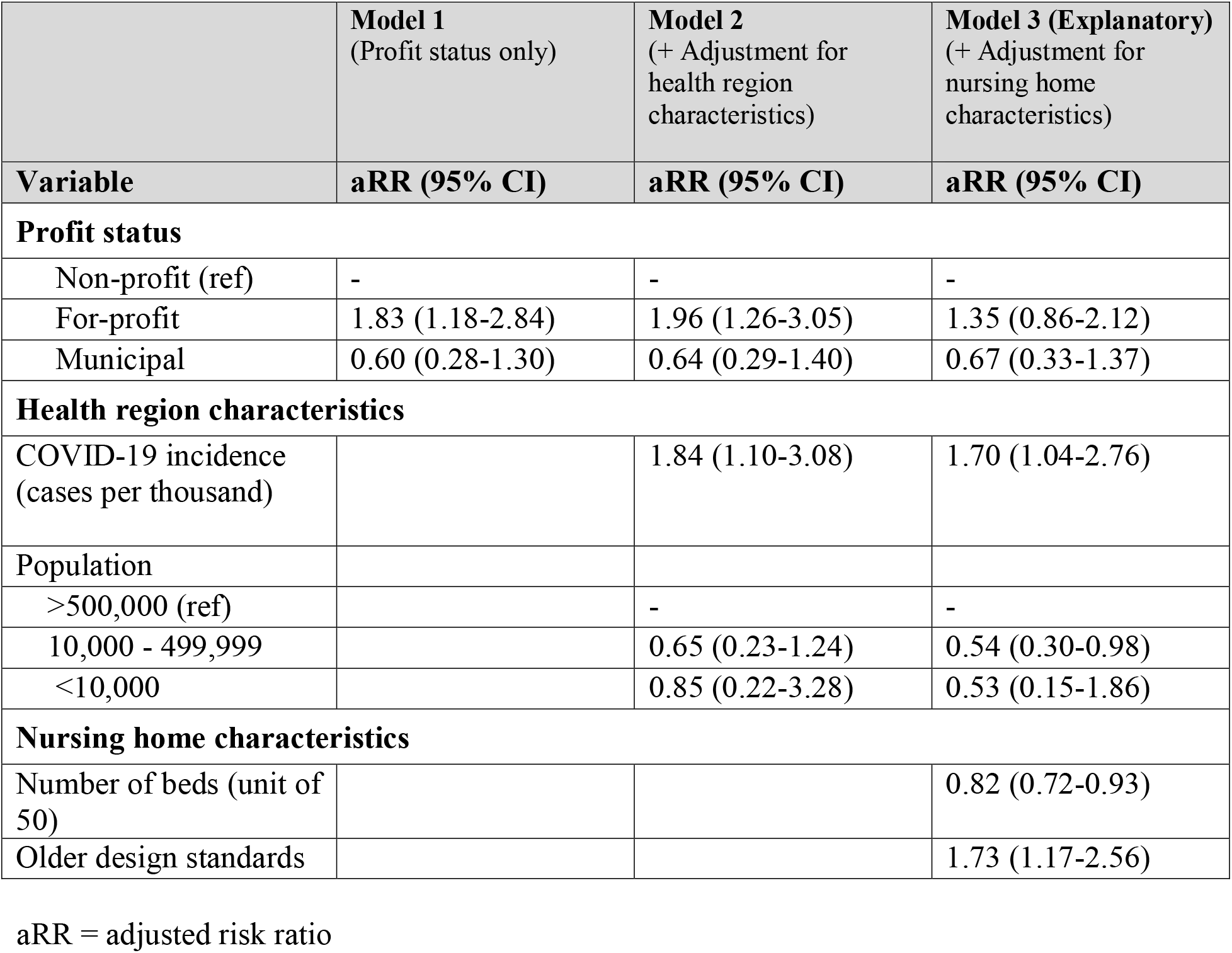
Size of Nursing Home COVID-19 Outbreaks by Profit Status.

|  | **Model 1**<br>(Profit status only) | **Model 2**<br>(+ Adjustment for<br>health region<br>characteristics) | **Model 3 (Explanatory)**<br>(+ Adjustment for<br>nursing home<br>characteristics) |
| --- | --- | --- | --- |
| **Variable** | **aRR (95% CI)** | **aRR (95% CI)** | **aRR (95% CI)** |
| **Profit status** |  |  |  |
| Non-profit (ref) | - | - | - |
| For-profit | 1.83 (1.18-2.84) | 1.96 (1.26-3.05) | 1.35 (0.86-2.12) |
| Municipal | 0.60 (0.28-1.30) | 0.64 (0.29-1.40) | 0.67 (0.33-1.37) |
| **Health region characteristics** |  |  |  |
| COVID-19 incidence<br>(cases per thousand) |  | 1.84 (1.10-3.08) | 1.70 (1.04-2.76) |
| Population |  |  |  |
| >500,000 (ref) |  | - | - |
| 10,000 - 499,999 |  | 0.65 (0.23-1.24) | 0.54 (0.30-0.98) |
| <10,000 |  | 0.85 (0.22-3.28) | 0.53 (0.15-1.86) |
| **Nursing home characteristics** |  |  |  |
| Number of beds (unit of<br>50) |  |  | 0.82 (0.72-0.93) |
| Older design standards |  |  | 1.73 (1.17-2.56) |

aRR = adjusted risk ratio

### Number of COVID-19 resident deaths in a nursing home outbreak

Among nursing homes with a COVID-19 outbreak, on average 6.5% of all residents in for-profit homes died of COVID-19, whereas on average 5.5% and 1.7% of all residents in non-profit and municipal homes died of COVID-19, respectively (Table 2). Seven of the ten homes with the highest death rates were for-profit-homes with older design standards (Figure 2). Quasi-Poisson regression modelling demonstrated that for-profit status was associated with the total number of nursing home resident COVID-19 deaths in the health region characteristics-adjusted model (aRR, 1.78; 95% CI 1.03-3.07) but not the unadjusted model (RR, 1.67; 95% CI 0.99-2.73) (Table 5). In the fully adjusted explanatory model, the relationship with for-profit status was attenuated (aRR, 1.19; 95% CI 0.70-2.01), and older nursing home design standards (aRR, 1.89; 95% CI 1.18-3.03) and number of beds (aRR = 0.80 (0.68-0.93), per 50 beds) were significantly associated with the risk of the total number of nursing home resident COVID-19 deaths.

**Figure 2:**
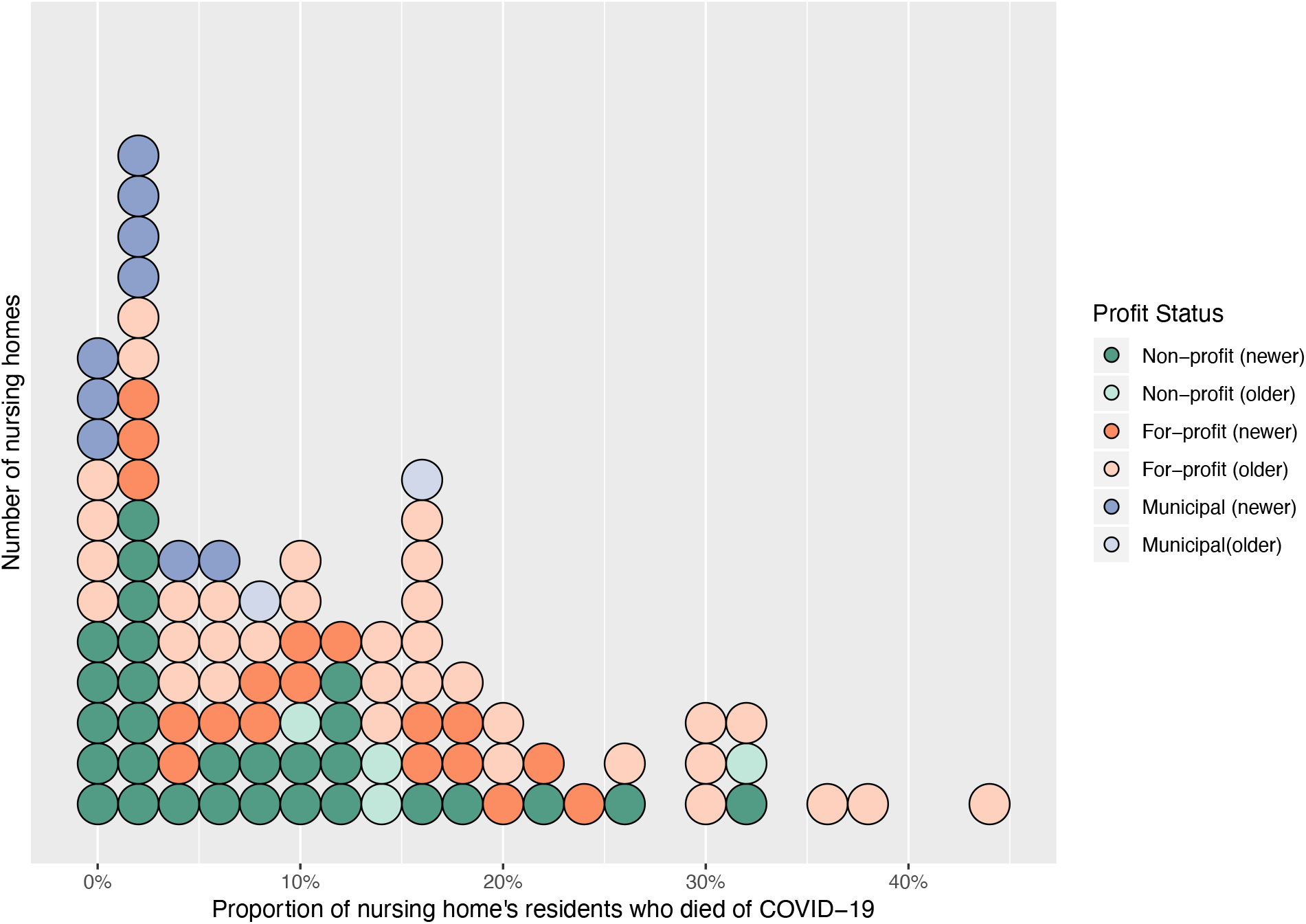
Distribution of the Number of COVID-19 Resident Deaths in a Nursing Home Outbreak by Profit Status.

**Table 5:**
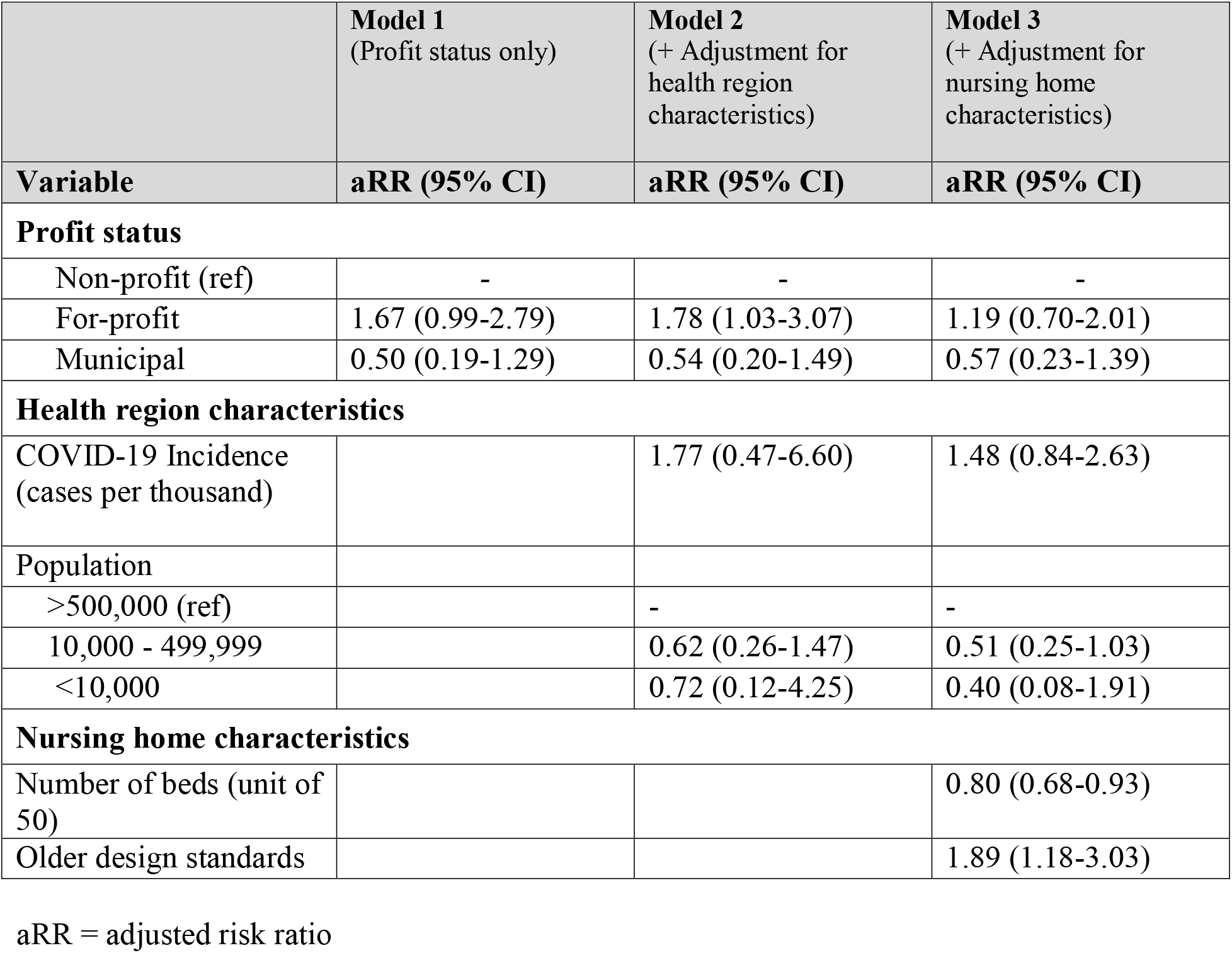
Number of Nursing Home Resident COVID-19 Deaths by Profit Status.

|  | **Model 1**<br>(Profit status only) | **Model 2**<br>(+ Adjustment for<br>health region<br>characteristics) | **Model 3**<br>(+ Adjustment for<br>nursing home<br>characteristics) |
| --- | --- | --- | --- |
| **Variable** | **aRR (95% CI)** | **aRR (95% CI)** | **aRR (95% CI)** |
| **Profit status** |  |  |  |
| Non-profit (ref) | - | - | - |
| For-profit | 1.67 (0.99-2.79) | 1.78 (1.03-3.07) | 1.19 (0.70-2.01) |
| Municipal | 0.50 (0.19-1.29) | 0.54 (0.20-1.49) | 0.57 (0.23-1.39) |
| **Health region characteristics** |  |  |  |
| COVID-19 Incidence<br>(cases per thousand) |  | 1.77 (0.47-6.60) | 1.48 (0.84-2.63) |
| Population |  |  |  |
| >500,000 (ref) |  | - | - |
| 10,000 - 499,999 |  | 0.62 (0.26-1.47) | 0.51 (0.25-1.03) |
| <10,000 |  | 0.72 (0.12-4.25) | 0.40 (0.08-1.91) |
| **Nursing home characteristics** |  |  |  |
| Number of beds (unit of<br>50) |  |  | 0.80 (0.68-0.93) |
| Older design standards |  |  | 1.89 (1.18-3.03) |

aRR = adjusted risk ratio

## Interpretation

In this study of all 623 nursing homes in Ontario, Canada we found that the odds of a COVID-19 nursing home outbreak was associated with the incidence of COVID-19 in the health region surrounding the nursing home and the total number of beds, but not for-profit status. Among nursing homes with a confirmed COVID-19 outbreak, for-profit status was associated with a 1.96-fold (95% CI 1.26-3.05) increase in outbreak size and a 1.78-fold (95% CI 1.03-3.07) increase in the number of COVID-19 resident deaths, after adjusting for health region characteristics and compared to non-profit homes. All comparisons favored municipal homes (Appendix 3).

The significant association between the risk of a nursing home outbreak and the incidence of COVID-19 in the surrounding health region is consistent with emerging literature demonstrating that nursing home staff are important vectors for SARS-CoV-2 transmission (4, 31). During the COVID-19 pandemic, most nursing homes have become relatively closed environments because of restrictions on visitors and resident transfers. This has meant that through no fault of their own, infected health care workers are the probable source of many nursing home outbreaks (32). An earlier study of COVID-19 outbreaks in Ontario nursing homes reported that staff infection was a significant predictor of future resident deaths (31). Screening protocols may be missing infected staff who are asymptomatic or minimally symptomatic, while low wages and scarce sick benefits mean that others may be working while ill (6, 7, 33). Many staff are also employed part-time and work at multiple healthcare facilities, something that has been linked to the transmission of COVID-19 between nursing homes (4, 34). Our additional observation associating the number of beds in a nursing home and the risk of an outbreak may be related to the fact that larger homes require more staff, thereby increasing the number of potential vectors for infection (35).

Our findings linking for-profit status with both the number of resident cases and deaths within nursing home COVID-19 outbreaks appears to be mediated, in large part, by the higher proportion of outdated design standards (which meet or fall below 1972 standards) in for-profit homes leading to more widespread transmission of COVID-19. Our hierarchical model revealed that associations favouring non-profit and municipal homes were attenuated when accounting for the age of a home’s design standards. Newer design standards provide for larger and more private room accommodations as well as less crowded and more self-contained common spaces which, beyond promoting quality of life, are designed to promote infection prevention and control (27).

The evolving COVID-19 crisis in Canada’s nursing homes has already led to sweeping calls for reforms to long-term care, including removing private for-profit businesses from the sector (6, 36). Our findings suggest that the incidence of COVID-19 in the health region surrounding a nursing home and the size of the home—but not for-profit status—are important risk factors for seeding COVID-19 nursing home outbreaks, whereas for-profit status (with for-profit homes more commonly having outdated design standards) is an important risk factor for transmission of COVID-19 after a home has been infected. Further, it is important to recognize that some, not all, for-profit homes have worse COVID-19 outcomes, as it seems to be those with older design standards likely indicating a failure to upgrade and modernize facilities. With governments like Ontario’s already committing to independent commissions and inquiries into their long-term care systems, it is important that policy recommendations and changes consider all root causes of the present crisis (37).

The study is limited by a lack of individual-level data on the sociodemographic and clinical characteristics of nursing home residents. Given the centralized admission process for long-term care in the Province of Ontario, we do not expect substantial differences in residents case-mix between for-profit, non-profit and municipal homes. Like other sources of data being rapidly collected during the COVID-19 pandemic, data from the Long-Term Care Inspections Branch was not independently validated, and there is potential for incompleteness. Since many nursing home outbreaks are still ongoing, there is also the possibility of right censoring of data; this limited our ability to study outbreaks which currently only involve staff but have the potential to spread to residents. We also could not account for changing provincial policies in infection prevention and control practices; however, these would not be expected to differentially impact for-profit homes. Finally, we did not account for SARS-CoV-2 testing rates, but COVID-19 case fatality rates between for-profit, non-profit and municipal homes were all similar suggesting similar levels of case identification.

In conclusion, we document that the risk of a COVID-19 nursing home outbreak was related to the COVID-19 infection rate in the health region surrounding the home and its total number of beds rather than for-profit status. We did find evidence that for-profit nursing homes have larger COVID-19 outbreaks and more COVID-19 resident deaths, compared to non-profit and municipal homes, and that this finding was mediated by the higher number of for-profit homes with outdated design standards. The COVID-19 pandemic has laid bare long-standing issues in how nursing homes are financed, operated and regulated (38). As health systems scramble to prepare nursing homes for successive waves of the COVID-19 pandemic and others search for accountability and solutions to the crisis in long-term care, it is important to examine all potential explanations for observed differences in COVID-19 outcomes across nursing homes.

## Data Availability

The underlying analytic code are available from the authors upon request (e-mail), understanding that the computer programs may rely upon coding templates or macros that are unique and are therefore either inaccessible or may require modification.

## Acknowledgements

We gratefully acknowledge the support of Michael Hillmer, Kamil Malikov and Sping Wang from the Ontario Ministry of Health’s Capacity Planning and Analytics Division for assistance with data acquisition, interpretation and analysis. Nathan M. Stall is supported by the Department of Medicine’s Eliot Phillipson Clinician-Scientist Training Program and the Clinician Investigator Program at the University of Toronto, and the Vanier Canada Graduate Scholarship. Paula A. Rochon holds the Retired Teachers of Ontario/ERO Chair in Geriatric Medicine at the University of Toronto. Andrew P. Costa holds the Schlegel Chair in Clinical Epidemiology and Aging at McMaster University.

## References

1. Barnett ML, Grabowski DC. Nursing Homes Are Ground Zero for COVID-19 Pandemic 2020 [Available from: https://jamanetwork.com/channels/health-forum/fullarticle/2763666.

2. Hsu AT, Lane N, Sinha SK, Dunning J, Dhuper M, Kahiel Z, et al. Impact of COVID-19 on residents of Canada’s long-term care homes - ongoing challenges and policy responses: International Long Term Care Policy Network,; 2020 [updated May 10, 2020. Available from: https://ltccovid.org/wp-content/uploads/2020/05/LTCcovid-country-reports_Canada_Hsu-et-al_May-10-2020.pdf.

3. D’Adamo H, Yoshikawa T, Ouslander JG. Coronavirus Disease 2019 in Geriatrics and Long-Term Care: The ABCDs of COVID-19. J Am Geriatr Soc. 2020.

4. McMichael TM, Currie DW, Clark S, Pogosjans S, Kay M, Schwartz NG, et al. Epidemiology of Covid-19 in a Long-Term Care Facility in King County, Washington. N Engl J Med. 2020.

5. Ouslander JG. Coronavirus Disease19 in Geriatrics and Long-Term Care: An Update. J Am Geriatr Soc. 2020.

6. Holroyd-Leduc JM, Laupacis A. Continuing care and COVID-19: a Canadian tragedy that must not be allowed to happen again. Cmaj. 2020.

7. Jabbar A, Raza D. Let’s keep profit out of long-term care [updated May 6, 2020. Available from: https://www.thestar.com/opinion/contributors/2020/05/06/lets-keep-profit-out-of-long-term-care.html.

8. Ronald LA, McGregor MJ, Harrington C, Pollock A, Lexchin J. Observational Evidence of For-Profit Delivery and Inferior Nursing Home Care: When Is There Enough Evidence for Policy Change? PLoS Med. 2016;13(4):e1001995.

9. Comondore VR, Devereaux PJ, Zhou Q, Stone SB, Busse JW, Ravindran NC, et al. Quality of care in for-profit and not-for-profit nursing homes: systematic review and meta-analysis. BMJ. 2009;339:b2732.

10. Hillmer MP, Wodchis WP, Gill SS, Anderson GM, Rochon PA. Nursing home profit status and quality of care: is there any evidence of an association? Med Care Res Rev. 2005;62(2):139–66.

11. McGregor MJ, Cohen M, McGrail K, Broemeling AM, Adler RN, Schulzer M, et al. Staffing levels in not-for-profit and for-profit long-term care facilities: does type of ownership matter? CMAJ. 2005;172(5):645–9.

12. Harrington C, Olney B, Carrillo H, Kang T. Nurse staffing and deficiencies in the largest for-profit nursing home chains and chains owned by private equity companies. Health Serv Res. 2012;47(1 Pt 1):106–28.

13. Hsu AT, Berta W, Coyte PC, Laporte A. Staffing in Ontario’s Long-Term Care Homes: Differences by Profit Status and Chain Ownership. Can J Aging. 2016;35(2):175–89.

14. McGregor MJ, Cohen M, Stocks-Rankin CR, Cox MB, Salomons K, McGrail KM, et al. Complaints in for-profit, non-profit and public nursing homes in two Canadian provinces. Open Med. 2011;5(4):e183–92.

15. Tanuseputro P, Chalifoux M, Bennett C, Gruneir A, Bronskill SE, Walker P, et al. Hospitalization and Mortality Rates in Long-Term Care Facilities: Does For-Profit Status Matter? J Am Med Dir Assoc. 2015;16(10):874–83.

16. Hirth RA, Grabowski DC, Feng Z, Rahman M, Mor V. Effect of nursing home ownership on hospitalization of long-stay residents: an instrumental variables approach. Int J Health Care Finance Econ. 2014; 14(1):1–18.

17. McGregor MJ, Abu-Laban RB, Ronald LA, McGrail KM, Andrusiek D, Baumbusch J, et al. Nursing home characteristics associated with resident transfers to emergency departments. Can J Aging. 2014;33(1):38–48.

18. McGregor MJ, Tate RB, McGrail KM, Ronald LA, Broemeling AM, Cohen M. Care outcomes in long-term care facilities in British Columbia, Canada. Does ownership matter? Med Care. 2006;44(10):929–35.

19. Grabowski DC, Feng Z, Hirth R, Rahman M, Mor V. Effect of nursing home ownership on the quality of post-acute care: an instrumental variables approach. J Health Econ. 2013;32(1): 12–21.

20. Damian J, Pastor-Barriuso R, Garcia-Lopez FJ, Ruigomez A, Martinez-Martin P, de Pedro-Cuesta J. Facility ownership and mortality among older adults residing in care homes. PLoS One. 2019;14(3):e0197789.

21. Castle NG, Wagner LM, Ferguson-Rome JC, Men A, Handler SM. Nursing home deficiency citations for infection control. Am J Infect Control. 2011;39(4):263–9.

22. Castle N, Wagner L, Ferguson J, Handler S. Hand hygiene deficiency citations in nursing homes. J Appl Gerontol. 2014;33(1):24–50.

23. Ontario Ministry of Health. COVID-19 Outbreak Guidance for Long-Term Care Homes (LTCH) 2020 [updated April 15, 2020. Available from: http://www.health.gov.on.ca/en/pro/programs/publichealth/coronavirus/docs/LTCH_outbreak_guidance.pdf.

24. Benchimol EI, Smeeth L, Guttmann A, Harron K, Moher D, Petersen I, et al. The REporting of studies Conducted using Observational Routinely-collected health Data (RECORD) statement. PLoS Med. 2015;12(10):e1001885.

25. von Elm E, Altman DG, Egger M, Pocock SJ, Gotzsche PC, Vandenbroucke JP, et al. Strengthening the Reporting of Observational Studies in Epidemiology (STROBE) statement: guidelines for reporting observational studies. BMJ. 2007;335(7624):806–8.

26. Daly T. Dancing the Two-Step in Ontario’s Long-term Care Sector: More Deterrence-oriented Regulation = Ownership and Management Consolidation. Stud Polit Econ. 2015;95(1):29–58.

27. Ontario Ministry of Health and Long-Term Care. Long-Term Care Home Design Manual 2015 February 2015 [Available from: http://health.gov.on.ca/en/public/programs/ltc/docs/home_design_manual.pdf.

28. Public Health Ontario. iPHIS Resources 2020 [Available from: https://www.publichealthontario.ca/en/diseases-and-conditions/infectious-diseases/ccm/iphis.

29. Harrison XA. Using observation-level random effects to model overdispersion in count data in ecology and evolution. PeerJ. 2014;2:e616.

30. Austin PC, Merlo J. Intermediate and advanced topics in multilevel logistic regression analysis. Stat Med. 2017;36(20):3257–77.

31. Fisman D, Lapointe-Shaw L, Bogoch I, McCready J, Tuite A. Failing our Most Vulnerable: COVID-19 and Long-Term Care Facilities in Ontario. medRxiv. 2020:2020.04.14.20065557.

32. Chow EJ, Schwartz NG, Tobolowsky FA, Zacks RLT, Huntington-Frazier M, Reddy SC, et al. Symptom Screening at Illness Onset of Health Care Personnel With SARS-CoV-2 Infection in King County, Washington. JAMA. 2020.

33. Pollock AM, Clements L, Harding-Edgar L. Covid-19: why we need a national health and social care service. BMJ. 2020;369:m1465.

34. Van Houtven CH, DePasquale N, Coe NB. Essential Long-Term Care Workers Commonly Hold Second Jobs and Double-or Triple-Duty Caregiving Roles. J Am Geriatr Soc. 2020.

35. Baldwin R, Chenoweth L, Dela Rama M, Wang AY. Does size matter in aged care facilities? A literature review of the relationship between the number of facility beds and quality. Health Care Manage Rev. 2017;42(4):315–27.

36. Canadian Labour Congress. Lessons from a Pandemic: Union Recommendations for Transforming Long-Term Care in Canada 2020 [Available from: http://documents.clcctc.ca/sep/LongTermCare-Report-EN.pdf.

37. Ontario Ministry of Long-Term Care. Ontario Announces Independent Commission into Long-Term Care 2020 [updated May 19, 2020. Available from: https://news.ontario.ca/mltc/en/2020/05/ontario-announces-independent-commission-into-long-term-care.html.

38. Grabowski DC, Mor V. Nursing Home Care in Crisis in the Wake of COVID-19. JAMA. 2020.

